# The development and usability testing of two arts-based knowledge translation tools for parents of children with functional constipation

**DOI:** 10.1101/2023.02.23.23286313

**Authors:** Alison Thompson, Anne Le, Lisa Hartling, Shannon D. Scott

## Abstract

Pediatric functional constipation (FC) is a common childhood problem that involves difficult or painful defecation and can be caused by a variety of different factors. In children, FC is often unrecognized and poorly treated, and has potential to cause abdominal pain, appetite suppression, loss of control over defecation, and family disruption. A recent interpretive description qualitative study found that parents who care for children with FC often experience a myriad of negative sentiments, including isolation and self-doubt. Furthermore, parents often have unanswered questions about the condition, particularly regarding the cause, symptoms, and treatment options. As such, more effective knowledge translation (KT) tools are needed to satisfy parental information needs.

The purpose of this research was to collaborate with parents to develop and test the usability of two animated KT tools (video and interactive infographic) on FC in children. Prototypes were co-developed with parents, and then evaluated by parents through usability testing in a large Alberta emergency department waiting room. Usability was assessed based on nine items with responses on a five-point Likert scale from 1=strongly disagree to 5=strongly agree. Overall, results were positive and the tools were highly rated across most usability items. Mean scores across usability items were 4.20 to 4.59 for the video and 3.73 to 4.30 for the infographic. The scores from the usability testing suggest arts-based digital tools are useful in sharing complex health information with parents about FC and provide meaningful guidance on how to improve KT tools to better reflect the needs of parents of children with FC.

## Introduction

Pediatric functional constipation (FC) is a common condition that impacts approximately 3% of children worldwide. It is characterized by difficult or painful passage of stools and can be caused by a variety of different factors (1). In children, FC should be addressed and managed immediately as inadequately treated FC can lead to issues such as abdominal pain, appetite suppression, loss of control over defecation, and family disruption (2, 3). Unfortunately, it has been shown that FC is often unrecognized and poorly treated (2). This can have significant impacts on families, with potential to cause frustration, confusion, and anger in both parents and children (2, 4).

In a recent interpretive description qualitative study, we found that parents who care for children with functional constipation often express feeling as if they are “living in the shadows” (5, p4). That is, parents described receiving negative reactions from other parents when discussing the “inappropriate” topics of defecation and incontinence, which as a result, deters them from openly discussing their child’s condition. Parents also described feeling that their concerns were overblown and dismissed by health care professionals, leading to self-doubt. Finally, parents with children who have functional constipation often have unanswered questions about the condition, particularly regarding the cause, symptoms, and treatment options (5, 6). This demonstrates the need to provide parents with accurate information and support that will enable them to make educated decisions about their child’s care.

Previous research has demonstrated the positive impacts of working with end-users of health information, such as parents and other caregivers, to develop knowledge translation (KT) tools (7-13). Such collaborations have resulted in tools that are relevant and meet the information needs of the appropriate stakeholders. More specifically, arts- and narrative-based KT tools have been proven to be effective sources of communication, translating complex health information into engaging and understandable content for parents (7-13). This is particularly important for complicated health conditions such as FC, where family education is considered a fundamental component of treatment (5). To date, there have been few tools developed to provide parents with guidance on how to manage their child’s FC despite the large body of evidence indicating that parents require these resources. As such, we sought to co-develop with parents, two arts-based KT tools about FC in children and test the usability of the tools. This report provides the results of the usability testing of the two KT tools that were developed.

## Methods

A series of studies using multiple methods and involving parent engagement were undertaken to develop, refine, and evaluate an animation video and interactive infographic for pediatric FC. Research ethics approval was obtained from the University of Alberta Health Research Ethics Board (Edmonton, Alberta) [Pro00062904 and Pro00087548]. Operational approvals were obtained from the pediatric emergency department to conduct usability testing. Results for the animation video are published elsewhere as part of a doctoral dissertation (14) and will be summarized here with the results for the infographic.

### Compilation of Parents’ Narratives

Our KT tools were informed by semi-structured interviews from an interpretive description qualitative study (5) (**Appendix A**) and a mixed methods systematic review (6). Parents were recruited through social media posts shared on health and parenting groups and posters displayed at facilities frequently visited by families (Facebook, Twitter, sports centres, libraries, etc.). Parents were asked to share their experiences with having a child with functional constipation. A systematic review was conducted to synthesize current evidence about experiences and information needs of parents managing functional constipation. Detailed methods and results from the qualitative study and the systematic review are published elsewhere (5, 6).

### Prototype (Intervention) Development

Results from both the systematic review and qualitative study were used to inform the development of a video script and an infographic skeleton. Clinical information from Bottom Line Recommendations developed by TRanslating Emergency Knowledge for Kids (TREKK) were also included in the infographic (15). Researchers worked with creative teams including illustrators and graphic designers to develop the tool prototypes.

### Video

The English-language video is approximately 5 minutes long, narrated in the third person, and includes closed captioning. It outlines the story of a 5-year old child named Ari who was struggling with FC. The video includes information about prevalence, typical age of onset, common symptoms, what to expect during assessment, physiological cycle of worsening symptoms, how soiling accidents occur, treatment, and further resources. Screen captures of the video are included in **Appendix B**.

### Infographic

The interactive infographic was developed in the same format as other infographics in our suite of tools (https://www.echokt.ca/tools/). The style is unique to our research program and was developed over the course of several years (16).

Functioning similarly to a webpage, the infographic utilizes a scroll feature that allows parents to explore at their own pace. As the page is opened and loaded, images and page content appear on the screen through various animations (slide in, zoom in, fade in, etc.). Each section of the infographic is a different color to ensure the tool is engaging and eye-catching to the viewer, as well as to delineate between different content. The infographic first introduces the topic at hand (FC) and describes what FC is, listing out potential causes. This is followed by six other sections: Diagnosis, Treatment, Medication, Accidents, What To Do, and Resources. Screen captures from the infographic are included in **Appendix C**.

### Revisions

We used iterative processes to develop the script and skeleton and sought feedback from parents, health care providers (HCPs), and researchers. HCPs were asked to comment on the accuracy of information and evidence, usefulness, and whether they would recommend the tools to their patients. Parents from our Pediatric Parents’ Advisory Group (P-PAG) (16) were asked to provide feedback on the length, illustrations, and whether the information was relevant to them as parents. Research team meetings were held weekly to discuss the development of the tools.

### Surveys

Parents presenting with an ill child to a major pediatric emergency department (ED) in the Edmonton area were recruited to participate in an electronic, usability survey (**Appendix D**). Research Staff from the Pediatric Emergency Medicine Research group at the Stollery approached parents in the ED to determine interest and study eligibility. Usability testing for the video was conducted between November 24, 2020 to December 17, 2020. Usability testing for the infographic was provided between January 21, 2021 to February 2, 2021. Parents who agreed to participate in the study were provided with an iPad by the researcher and asked to complete a consent form. Study team members were available in the ED to provide technical assistance and answer questions as parents were completing the surveys. The usability survey included 9, 5-point Likert items that assessed: 1) usefulness, 2) aesthetics, 3) length, 4) relevance, and 5) future use. The usability survey was designed in-house based upon key elements identified by a systematic search of over 180 usability evaluations (17). Parents were also asked to provide their positive and negative opinions of the tool via two free text boxes.

### Data Analysis

Data was cleaned and analyzed using SPSS v.24. Descriptive statistics and measures of central tendency were generated for demographic questions. Likert responses were given a corresponding numerical score, with 5 being “Strongly Agree” and 1 being “Strongly Disagree” (18, 19). Means and standard deviations (SDs) were calculated for each usability item. Independent two tailed t-tests were used to determine if there was a significant difference in the mean usability scores of the two KT tools for each usability item. Open-ended survey data was analyzed thematically.

See **Appendix E** for an overview of the entire project timeline.

## Results

Sixty parents awaiting pediatric ED care completed the usability survey (30 parents participated in the infographic usability testing and 30 parents participated in the video usability testing). The demographic characteristics of the parents who participated are presented in **Table 1**.

**Table 1.** Demographic characteristics of parents who assessed the usability of the FC video and interactive infographic (video n=30; infographic n=30; combined n=60)

| Characteristic | Video<br>n (%) | Infographic<br>n (%) | Combined<br>n (%) |
| --- | --- | --- | --- |
| Sex |  |  |  |
| Female | 24 (80.0) | 23 (76.7) | 47 (78.3) |
| Male | 6 (20.0) | 7 (23.3) | 13 (21.7) |
| Ethnicity |  |  |  |
| African American/Canadian | 3 (10.0) | 1 (3.3) | 4 (6.7) |
| Asian | 2 (6.7) | 2 (6.7) | 4 (6.7) |
| First Nations | 2 (6.7) | 1 (3.3) | 3 (5.0) |
| Metis | 1 (3.3) | 2 (6.7) | 3 (5.0) |
| Middle Eastern or North African | 2 (3.3) | 0 (0.0) | 2 (3.3) |
| South Asian | 3 (10.0) | 1 (3.3) | 4 (6.7) |
| White/Caucasian | 15 (50.0) | 22 (73.7) | 37 (61.7) |
| Other (not listed) | 1 (3.3) | 0 (0.0) | 1 (1.7) |
| Prefer not to answer | 1 (3.3) | 1 (3.3) | 2 (3.3) |
| Age |  |  |  |
| Less than 20 | 0 (0.0) | 1 (3.3) | 1 (1.7) |
| 20-30 years | 4 (13.3) | 5 (16.7) | 9 (15.0) |
| 31-40 years | 20 (66.7) | 13 (43.4) | 33 (55.0) |
| 41-50 years | 6 (20.0) | 8 (26.7) | 14 (23.3) |
| 51-60 years | 0 (0.0) | 3 (10.0) | 3 (5.0) |
| Marital Status |  |  |  |
| Married/Partnered | 25 (83.3) | 22 (73.3) | 47 (78.3) |
| Single | 5 (16.7) | 8 (26.7) | 13 (21.7) |
| Education |  |  |  |
| Some high school | 3 (10.0) | 3 (10.0) | 6 (10.0) |
| High school diploma | 2 (6.7) | 4 (13.3) | 6 (10.0) |
| Some post-secondary | 3 (10.0) | 4 (13.3) | 7 (11.7) |
| Post-secondary certificate/diploma | 7 (23.3) | 8 (26.7) | 15 (25.0) |
| Post-secondary degree | 11 (36.7) | 8 (26.7) | 19 (31.7) |
| Graduate degree | 4 (13.3) | 3 (10.0) | 7 (11.7) |
| Household Income |  |  |  |
| Less than \$25,000 | 4 (13.3) | 1 (3.3) | 5 (8.3) |
| \$25,000-\$49,000 | 5 (16.7) | 4 (13.3) | 9 (15.0) |
| \$50,000-\$74,000 | 6 (20.0) | 5 (16.7) | 11 (18.3) |
| \$75,000-\$99,000 | 2 (6.7) | 5 (16.7) | 7 (11.7) |
| \$100,000-\$149,000 | 6 (20.0) | 4 (13.3) | 10 (16.7) |
| \$150,000 and over | 5 (16.7) | 6 (20.0) | 11 (18.3) |
| Prefer not to answer | 2 (6.7) | 5 (16.7) | 7 (11.7) |
| Geographical Area |  |  |  |
| City | 21 (70.0) | 24 (80.0) | 45 (75.0) |
| Farm | 1 (3.3) | 0 (0.0) | 1 (1.7) |
| Town | 1 (3.3) | 1 (3.3) | 2 (3.3) |
| Suburb | 6 (20.0) | 4 (13.3) | 10 (16.7) |
| Other | 1 (3.3) | 1 (3.3) | 2 (3.3) |
| Number of Children in the Family |  |  |  |
| 1 | 9 (30.0) | 6 (20.0) | 15 (25.0) |
| 2 | 10 (33.3) | 14 (46.7) | 24 (40.0) |
| 3 | 8 (26.7) | 7 (23.3) | 15 (25.0) |
| 4 | 3 (10.0) | 1 (3.3) | 4 (6.7) |
| 5+ | 0 (0.0) | 1 (3.3) | 1 (1.7) |
| Missing | 0 (0.0) | 1 (3.3) | 1 (1.7) |
| Times Visited ED |  |  |  |
| 1-5 Times | 23 (76.7) | 26 (86.7) | 49 (81.7) |
| 6+ Times | 7 (23.3) | 4 (13.3) | 11 (18.3) |

Overall, the tools were rated highly by parents with most selecting strongly agree and agree for the usability items (**Table 2**). The mean scores across usability items for the video ranged from 4.20 to 4.59 (on a 5-point scale with 4 indicating agree and 5 indicating strongly agree). Parents felt the video was useful and relevant to them. They found the video easy to use and felt it could be used without written instruction. Parents also found the length of the video appropriate and thought it was aesthetically pleasing. Parents agreed and strongly agreed that they would use the video in the future and that it would help them make decisions about their child’s health. Finally, when asked if they would recommend the video to a friend, parents agreed or strongly agreed.

**Table 2.** Means and standard deviation (SD) of participant responses to the usability survey

| **Usability Measures** | **Video**<br>**mean (SD)** | **Infographic**<br>**mean (SD)** | **Significance**<br>**(p-value)** |
| --- | --- | --- | --- |
| It is useful. | 4.47 (0.57) | 4.13 (0.68) | 0.045* |
| It provides information that is relevant to me as a parent. | 4.43 (0.63) | 4.03 (0.81) | 0.036* |
| It is simple to use. | 4.50 (0.51) | 4.30 (0.60) | 0.167 |
| I can use it without written instructions or additional help. | 4.40 (0.49) | 4.30 (0.65) | 0.507 |
| Its length is appropriate. | 4.23 (0.63) | 4.23 (0.68) | 1.00 |
| It is aesthetically pleasing (i.e. images, colours, etc.). | 4.30 (0.65) | 4.27 (0.69) | 0.848 |
| It helps me make decisions about my child's health. | 4.20 (0.61) | 3.87 (0.78) | 0.070 |
| I would use it in the future. | 4.20 (0.71) | 3.73 (0.78) | 0.019* |
| I would recommend it to a friend. | 4.27 (0.64) | 3.82 (0.72) | 0.016* |

\*p value of <0.05 is considered significant.

For the infographic, mean scores across usability items ranged from 3.73 to 4.30 (where 3 indicates neither agree nor disagree and 4 indicates agree). Parents felt that the infographic was useful and relevant to them as parents. When parents were asked whether the infographic was simple to use, parents agreed and strongly agreed and also felt that the tool could be used without written instructions or additional help.

Parents felt that the length of the infographic was appropriate and that it was aesthetically pleasing. However, parents’ scores were slightly lower when asked about potential future use of the infographic, whether they would use the infographic to make decisions about their child’s health, and if they would recommend the tool to a friend.

There was a statistically significant difference in the mean ratings for the video versus the infographic on four of the usability measures. Mean ratings for the video were significantly higher for the following items: it is useful, it provides information that is relevant to me as a parent, I would use it in the future, and I would recommend it to a friend. There were no statistically significant differences in the mean ratings for simplicity, ability to use without written instructions of help, length, aesthetics, and helpfulness for decision making for the video versus the infographic.

There were a couple open text responses describing the video as “a bit too long” in the negative aspects, however the responses for positive aspects included many comments about how the video was “easy to understand”, “clear” and “informative” as well as “visually appealing”. Open text responses for the infographic included many comments about how the tool was simple to use and easy to understand. However, the comments did not elucidate why parents were hesitant to use or recommend the infographic as a decision-making tool in the future. As a result of these comments and the high rating of both tools, no revisions were made to the tools following usability testing.

## Conclusions

Through stakeholder engagement, our research group developed and tested an animated video and an interactive infographic for parents of children with FC. Usability testing demonstrated that overall, parents liked the tools, rating them above average for all usability items. In particular, parents found the tools to be useful, simple to use, relevant, aesthetically pleasing, and that the length of both tools was appropriate.

Parental usability scores for the infographic were slightly lower than the video. Despite these findings, parents recommended both tools for future use and would recommend both tools to their friends.

The scores from our usability tests suggest that arts-based digital KT tools are useful in sharing complex health information with parents and provide meaningful guidance on how to improve these tools to better reflect the needs of parents.

### The tools can be found here

<u>echokt.ca/functional-constipation</u>

Note: Our KT tools are assessed for alignment with current, best-available evidence every two years. If recommendations have changed, appropriate modifications are made to our tools to ensure that they are up-to-date (20).

## Other Outputs from this Project

### Research Papers

Thompson AP. Understanding parents’ experiences and information needs to inform a digital knowledge translation tool about pediatric functional constipation: University of Alberta; 2021. doi:10.7939/r3-3gqn-tf74

Thompson AP, MacDonald SE, Scott SD, Wine E. Understanding parents’ experiences when caring for a child with functional constipation: Interpretive description study. JMIR Pediatrics and Parenting. 2021;4(1): e2485. doi: 10.2196/24851

Thompson AP, Wine E, MacDonald SE, Campbell A, Scott SD. Parents’ Experiences and Information Needs While Caring for a Child with Functional Constipation: A Systematic Review. Clinical Pediatrics. 2021;60(3): 154-69. doi: 10.1177/0009922820964457

Thompson AP, MacDonald SE, Wine E, Scott SD. An Evaluation of Parents’ Experiences of Patient Engagement in Research to Develop a Digital Knowledge Translation Tool: Protocol for a Multi-Method Study. JMIR Res Protoc. 2020, 9(8): e19108. doi: 10.2196/19108.

### Presentations & Research Conferences

Thompson, A., Wine, E., MacDonald, S., & Scott, S.D. A systematic review of parents’ experiences and information needs related to pediatric functional constipation. Women and Children’s Health Research Institute Research Day. Edmonton, AB. November 6, 2019.

Thompson, A., MacDonald, S., Wine, E., & Scott, S.D. Living in the Shadows: A Qualitative Exploration of Parents’ Experiences Caring for a Child with Functional Constipation. Qualitative Health Research Conference, Vancouver, Canada. October 25-29th, 2019.

Thompson, A. & Scott, S.D. Development and Evaluation of a KT Tool for Parents with a Child with Functional Constipation. KT Canada Summer Institute, Toronto, Canada. June 26-28th 2017.

## Supporting information

Supplemental Table 1: Adapted Reporting Checklist for Multi-Method research

## Data Availability

All data produced in the present study are available upon reasonable request to the authors

https://www.echokt.ca/functional-constipation/

## Author Contributions

This work was conducted as a part of a larger research program led by Dr. Shannon D. Scott (SDS) and Dr. Lisa Hartling (LH), principal investigators (PIs) for **translating Evidence in Child Health to enhance Outcomes** (ECHO) Research and the **Alberta Research Centre for Health Evidence** (ARCHE), respectively. The development of the video arts-based knowledge translation (KT) tool for parents of children with functional constipation (FC) was the focus of Alison Thompson’s (AT) doctoral dissertation work at the University Alberta under the supervision of SDS.

AT designed the FC arts-based video and completed the evaluation of this tool with input from her supervisory committee which included SDS as her primary supervisor.

The usability data were analyzed by AT and Anne Le (AL) for the video and AL for the infographic.

SDS designed and supervised all aspects of development and evaluation of the FC infographic.

LH designed and provided input on all aspects of development and evaluation of the FC infographic.

All authors contributed to the writing of this technical report and provided substantial feedback.

## Acknowledgements

Given COVID-19 pandemic restrictions which prevented ECHO research staff from collecting the usability data, University of Alberta Pediatric Emergency Medicine research staff were contracted to collect usability data.

Dr Shannon MacDonald and Dr. Eytan Wine were members of AT’s doctoral dissertation committee. They advised on the qualitative study and the mixed studies systematic review that informed the development of these KT tools. They provided input on all aspects of the development and evaluation of the arts-based video.

## Funding

This work was funded by:

### Canadian Institutes of Health Research

- Scott, S.D. (co-PI), Hartling, L. (co-PI), Ali, S., Currie, G., Dyson, M., Fernandes, R., Fleck, B., Freedman, S., Jabbour, M., Johnson, D., Junker, A., Klassen, T.,Maynard, D., Newton, A., Plint, A., Richer, L., Robinson, J., Robson, K., Vandall-Walker, V. [all collaborators listed in alphabetical order]. (2016). Integrating evidence and parent engagement to optimize children’s healthcare. CIHR Foundation Scheme ($2,500,000). July 2016-June 2026.
- Studentship Funding (awarded to AT) also supported this project, as well as matched funds awarded to Scott and Hartling as part of the Translating emergency knowledge for kids (TREKK) project.

Infrastructure funding was provided by:

### Networks of Centres of Excellence

- Klassen, T., Hartling, L., Jabbour, M., Johnson, D., & Scott, S.D. (2015). Translating emergency knowledge for kids (TREKK). Networks of Centres of Excellence of Canada Knowledge Mobilization Renewal ($1,200,000). January 2016 – December 2019.

### Stollery Children’s Hospital Foundation and the Women’s and Children Health Research Institute

- Scott, S.D., Hartling, L. Distinguished Researcher Funding. (2018). Women and Children’s Health Research Institute & Stollery Children’s Hospital Foundation ($1,000,000). September 2018 – August 2023.

## Appendices

### Appendix A – Qualitative Interview

#### Project Title - Collaborating with Parents to Understand and Address Information Needs when Caring for a Child with Functional Constipation

1. Tell me about your experiences of having a child with constipation?
2. What did you understand about constipation when your child was first diagnosed?
  - What information were you provided with from health care providers when your child was first diagnosed? What, if anything, was helpful about this information?
  - When did they give you this information?
  - In what format did the health care providers give you this information? Written, verbal, demonstration, combination?
3. How would you have liked to receive this information?
4. How did this information influence your experience with your child’s illness?
5. If your friend’s child had constipation and she asked you for information, what would you teach her about it?
  - When would you give her this information?
6. Today, do you feel you have enough information about your child’s constipation?
  - If no, what would you like more information on?
7. How have your learning needs changed over time (comparison between time of diagnosis and now?)
  - Better access to information?
  - Relationship with health care personnel?
8. How has your confidence to manage your child’s illness changed over time? What has influenced your confidence level over time?
9. What was has been the hardest part of having a child with constipation? How has that changed over time?
10. What would you like health providers to know about your experiences of living with a child who has constipation?
11. What is important to you in terms of your child’s health outcomes?
12. How do you “define” when your child has a good day? E.g. Number of stools per day? No complaints of pain? No soiling? Child attending school or activities? Parents being able to go to work?

Thank you for your thoughtful feedback to my questions. Do you have any questions or concerns?

### Appendix B – Images from arts-based video about functional constipation (https://www.echokt.ca/functional-constipation/)

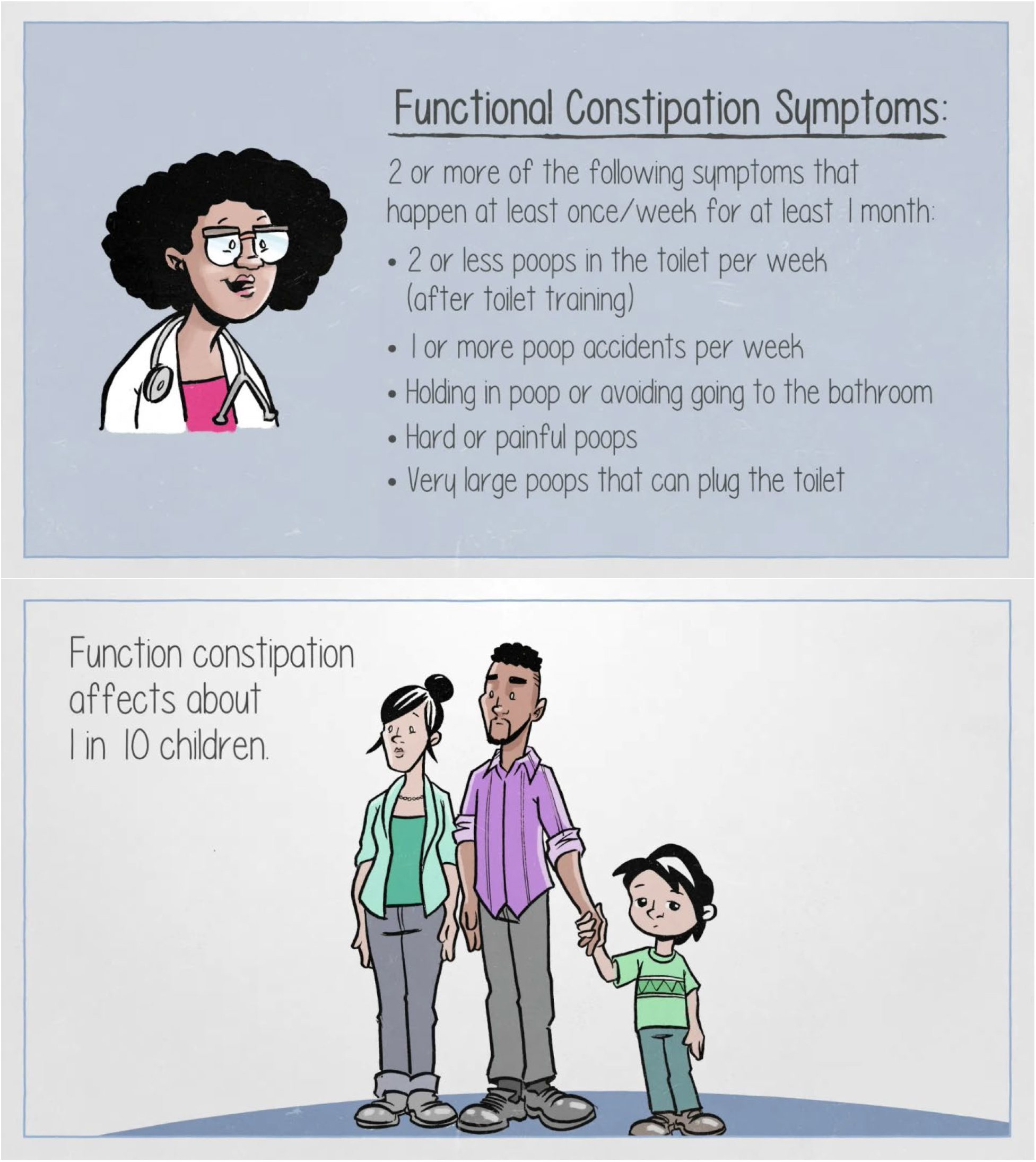

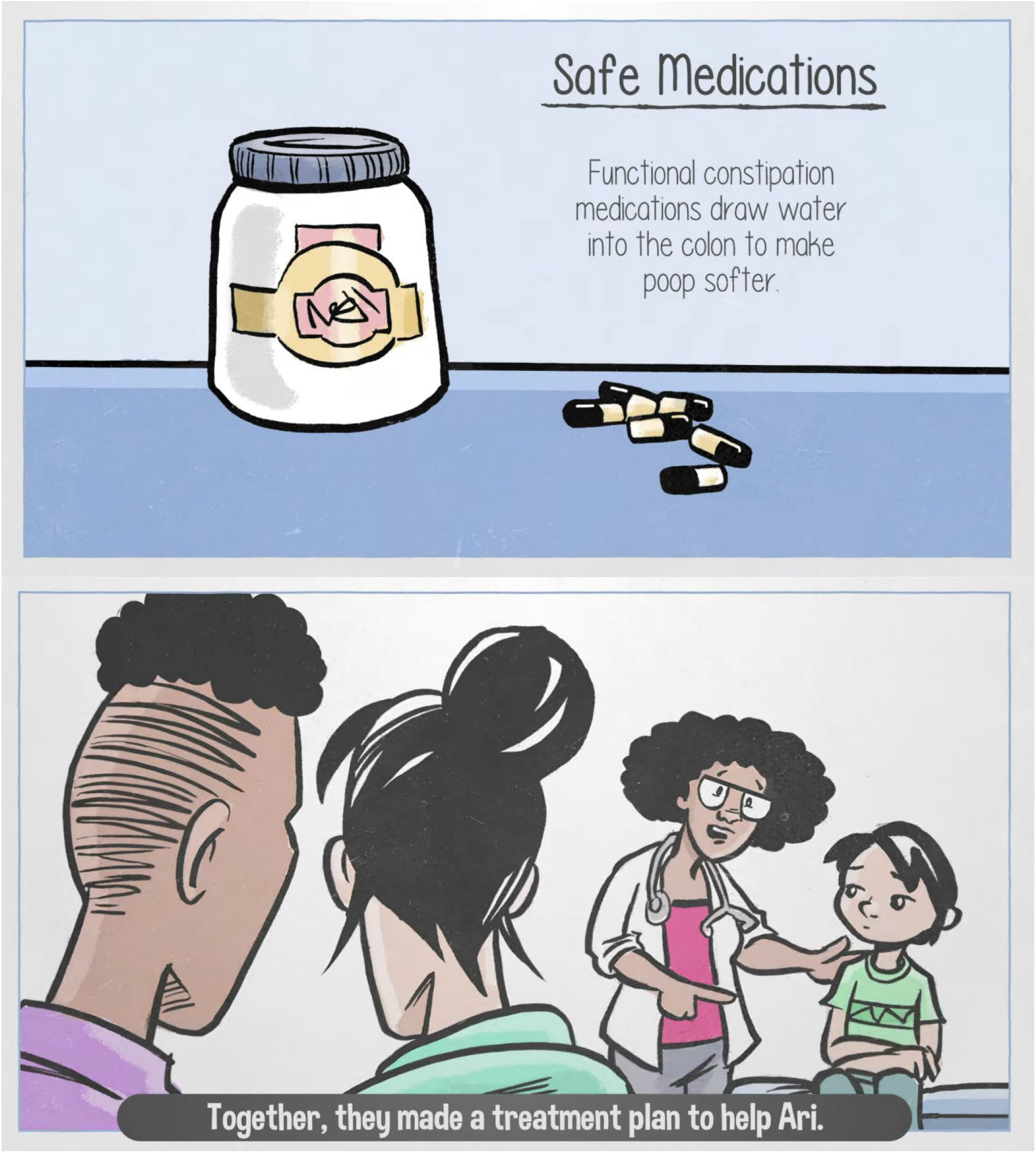

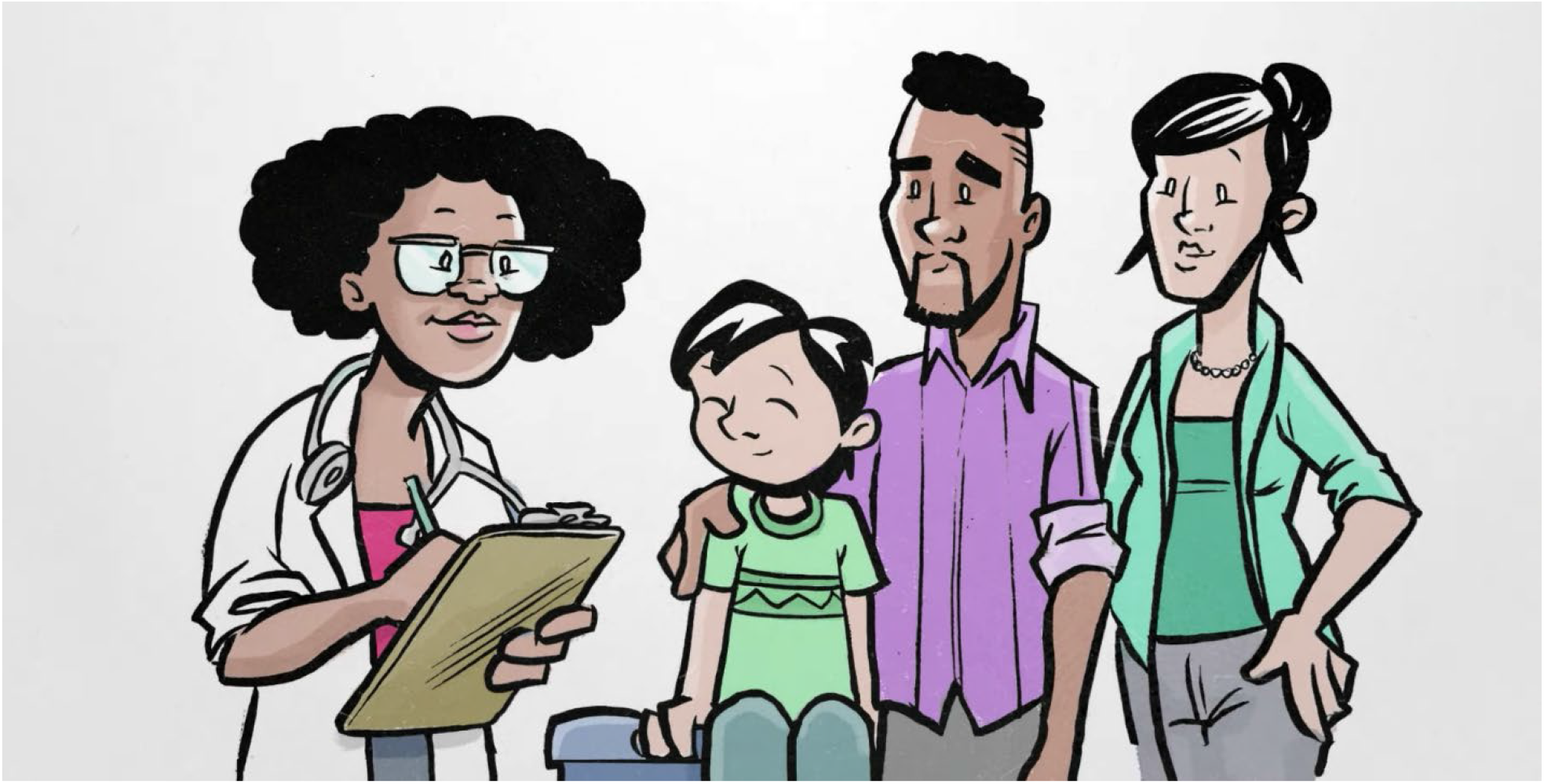

### Appendix C – Images from Infographic about functional constipation (https://www.echokt.ca/fc-infographic/)

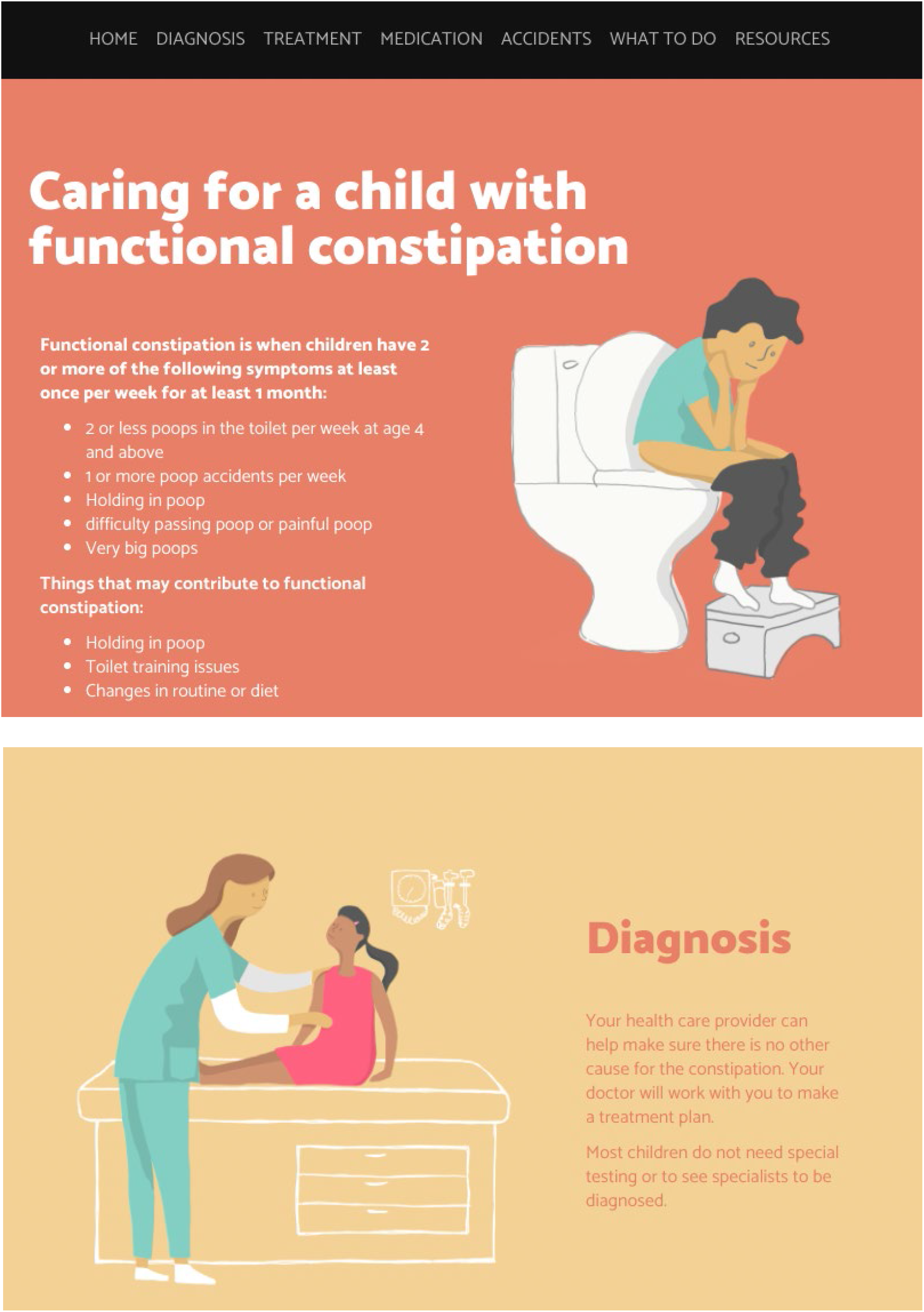

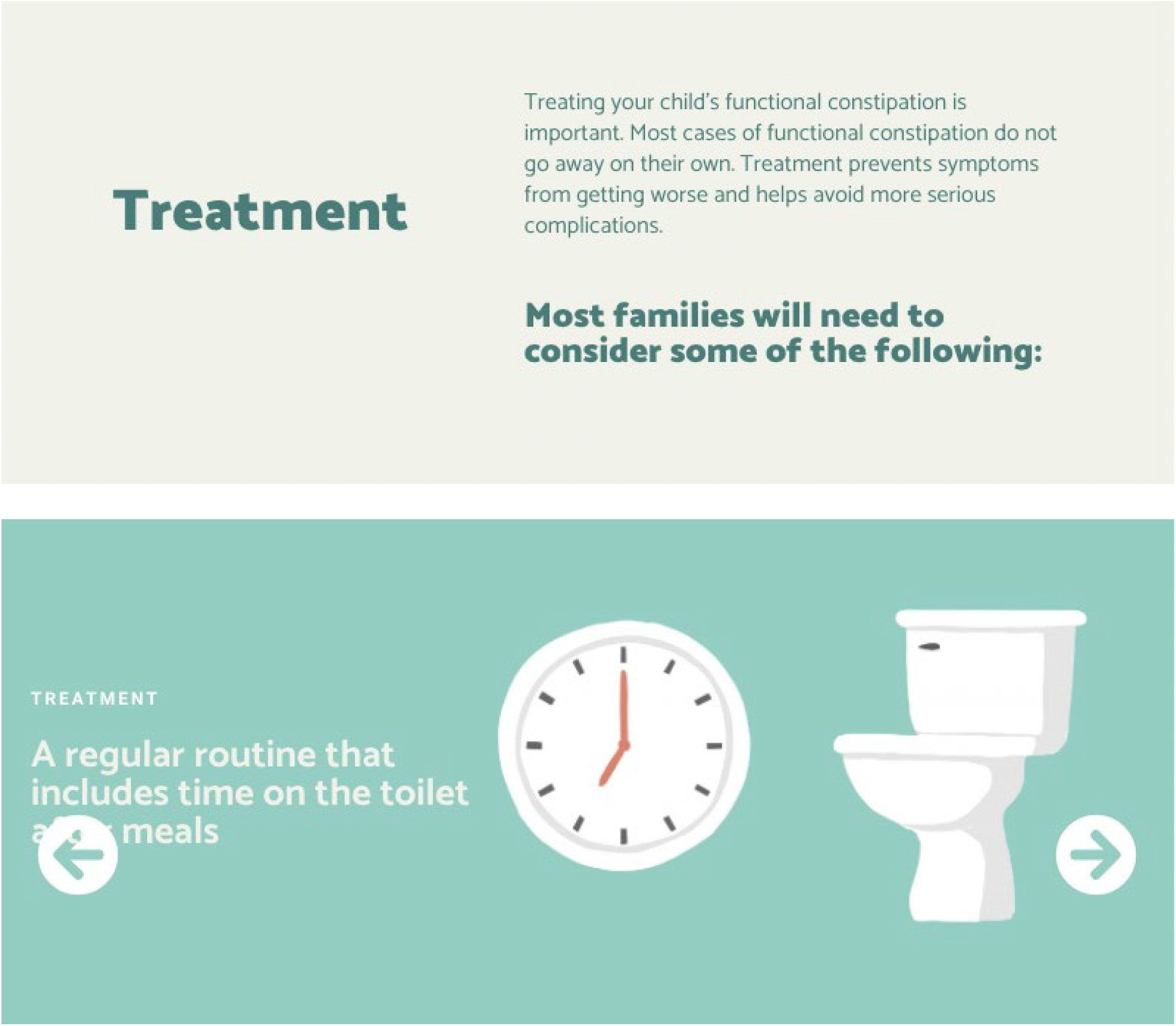

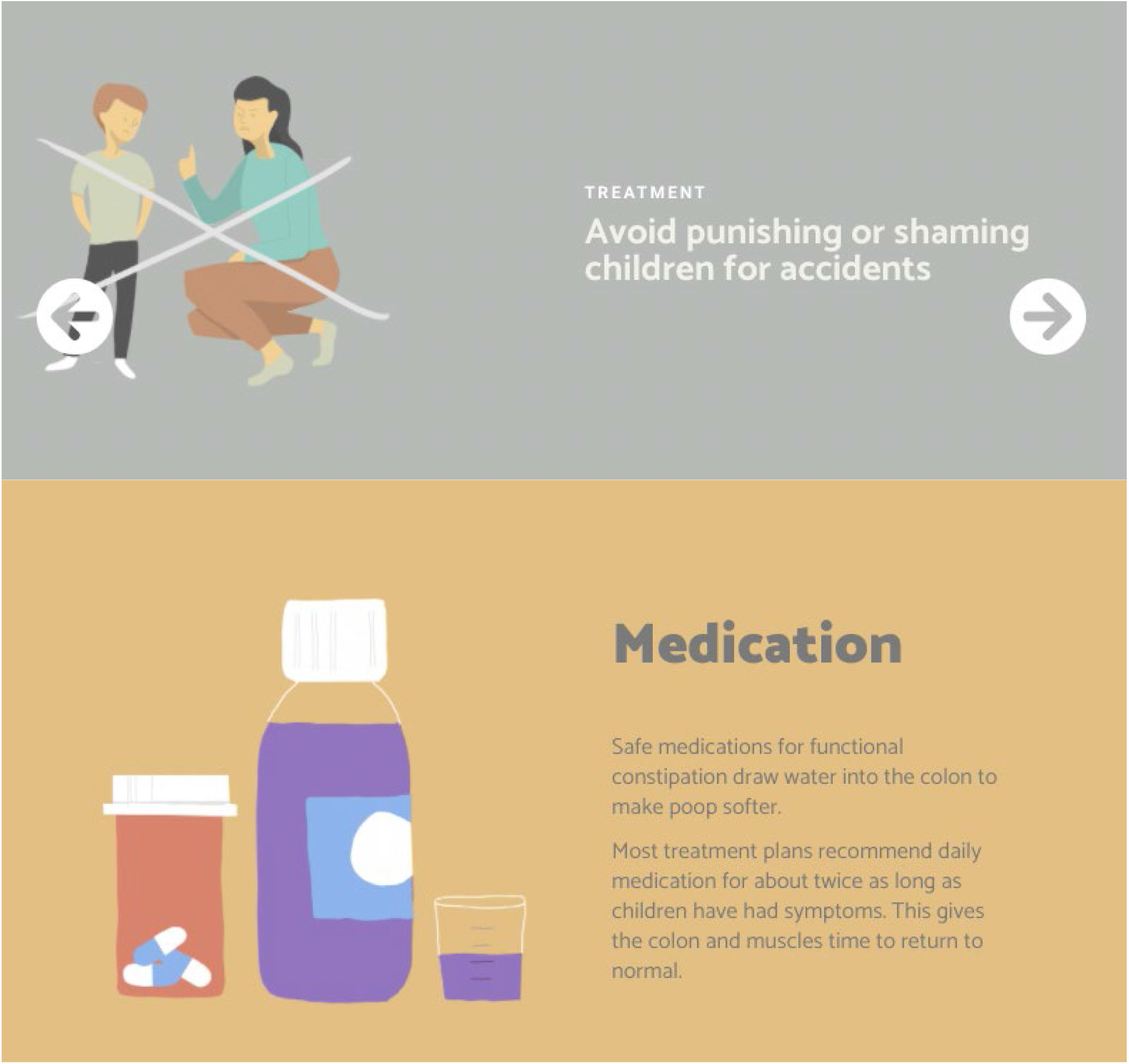

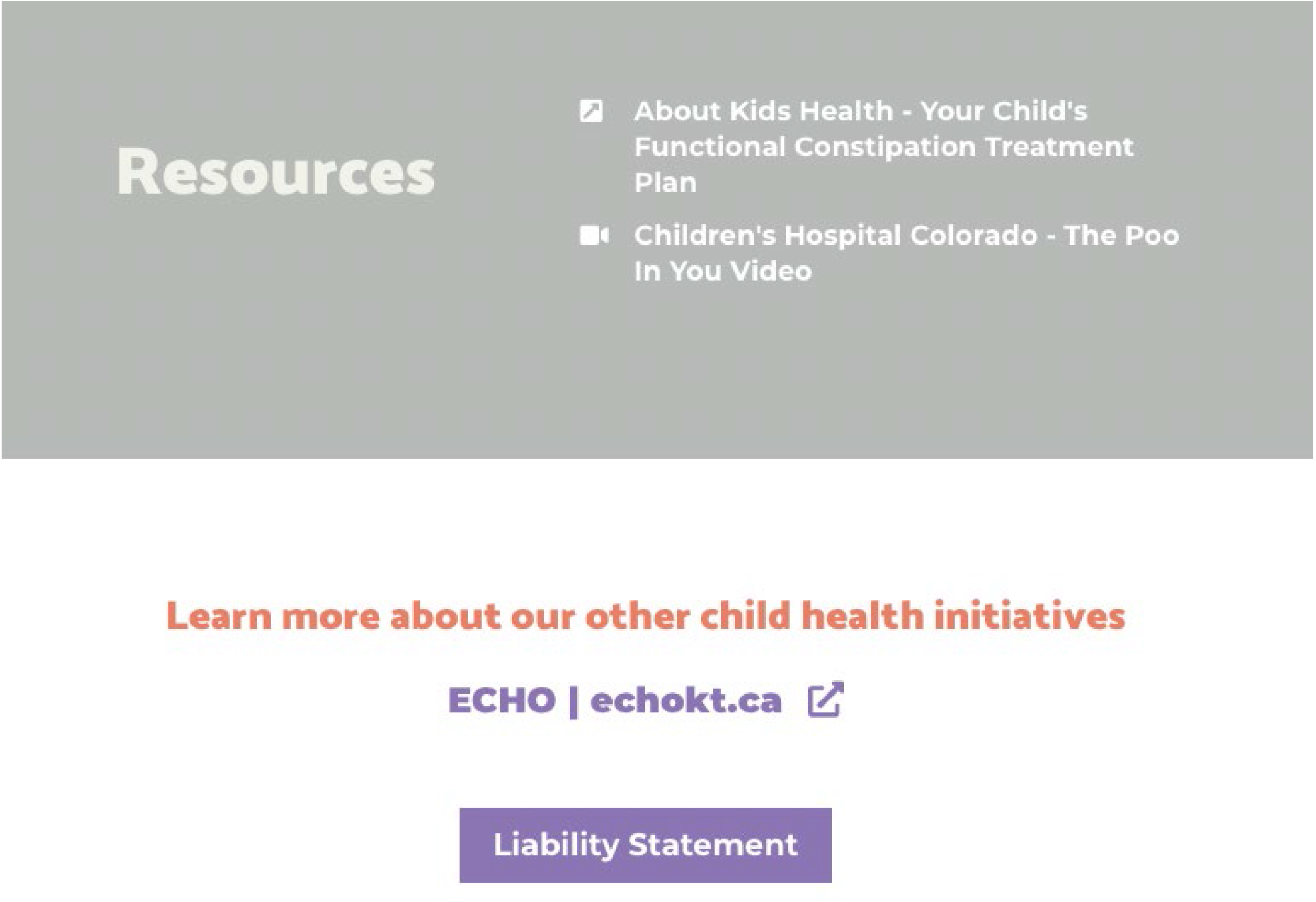

### Appendix D – Usability Survey

SECTION 1: Demographics

1) a. Which gender do you identify with most?
  □ Male
  □ Female
  □ Non-binary
  □ Two-spirit
  □ Other:
  □ Prefer not to answer
1) b. Would you describe yourself as transgender?
  □ Yes
  □ No
  □ Prefer not to answer
2) Which ethnicities best describes you? *Please select all that apply*.
  □ Asian
  □ African American or African Canadian
  □ Black
  □ First Nations
  □ Hispanic or Latino
  □ Métis
  □ Middle Eastern or North African
  □ South Asian
  □ Southeast Asian
  □ White or Caucasian
  □ Not listed:
  □ Prefer not to answer
3) What is your Age?
  □ Less than 20 years old
  □ 20-30 years
  □ 31-40 years
  □ 41-50 years
  □ 51 years and older
4) What is your Marital Status?
  □ Married/Partnered
  □ Single
5) What is your gross annual household income?
  □ Less than $25,000
  □ $25,000-$49,999
  □ $50,000-$74,999
  □ $75,000-$99,999
  □ $100,000-$149,999
  □ $150,000 and over
  □ Prefer not to answer
6) What is your highest level of education?
  □ Some high school
  □ High school diploma
  □ Some post-secondary
  □ Post-secondary certificate/diploma
  □ Post-secondary degree
  □ Graduate degree
  □ Other:
7) Where does your household live
  □ City
  □ Suburb
  □ Town
  □ Farm
  □ Other:
8) What is your relationship to the child that you have brought to the emergency department?
  □ Parent
  □ Grandparent
  □ Other family member
  □ Guardian
9) How many children do you have?
10) How old are your children?
11) How many times have you visited the emergency department with your children?
  □ 1-5 times
  □ 6+times
12) Have any of your children ever been admitted to the hospital?
  □ Yes
  □ No

SECTION 2: Assessment of attributes of the arts-based, digital tools

Note: items 1-9 are rated on a 5-point Likert scale from 1=strongly disagree to 5=strongly agree

1. It is useful.
2. It provides information that is relevant to me as a parent.
3. It is simple to use.
4. I can use it without written instructions or additional help.
5. Its length is appropriate.
6. It is aesthetically pleasing (i.e., images, colours, etc.).
7. It helps me to make decisions about my child’s health.
8. I would use it in the future.
9. I would recommend it to a friend.
10. List the most negative aspects: [open text]
11. List the most positive aspects: [open text]

### Appendix E – Project Timeline

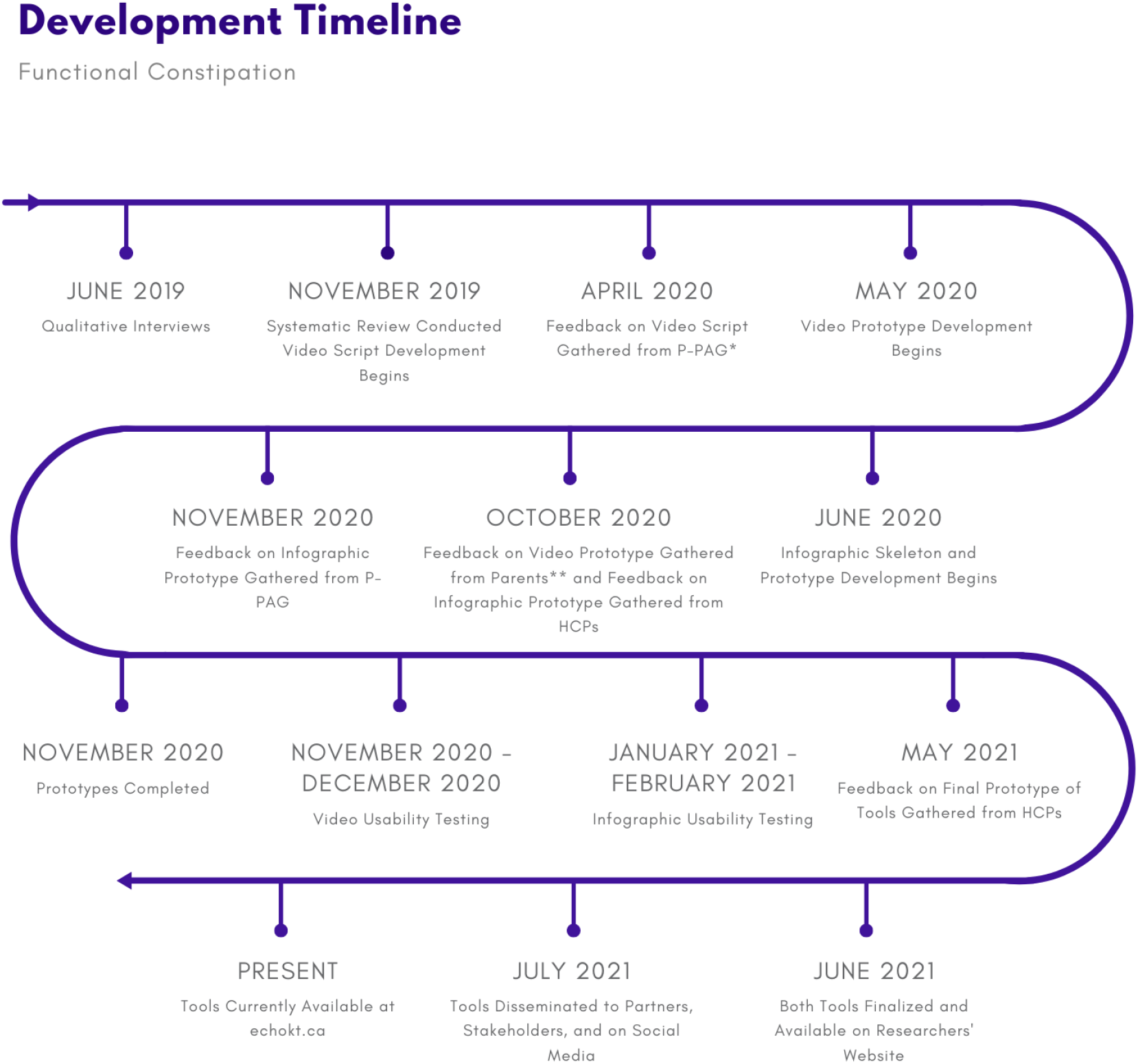

* P-PAG = Pediatric Parents’ Advisory Group (P-PAG)

** Refers to parents from Pediatric Parents’ Advisory Group (P-PAG) HCPs = Healthcare Providers

